# Outbreak dynamics of COVID-19 in Europe and the effect of travel restrictions

**DOI:** 10.1101/2020.04.18.20071035

**Authors:** Kevin Linka, Mathias Peirlinck, Francisco Sahli Costabal, Ellen Kuhl

**Affiliations:** Departments of Mechanical Engineering and Bioengineering, Stanford University, Stanford, California, United States; Department of Mechanical and Metallurgical Engineering, School of Engineering; Institute for Biological and Medical Engineering, Schools of Engineering, Medicine and Biological Sciences, Pontificia Universidad Catolica de Chile, Santiago, Chile

**Keywords:** Coronavirus, COVID-19, epidemiology, SEIR model, outbreak dynamics, outbreak control

## Abstract

For the first time in history, on March 17,2020, the European Union closed all its external borders to contain the spreading of the coronavirus 2019, COVID-19. Throughout two past months, governments around the world have implemented massive travel restrictions and border control to mitigate the outbreak of this global pandemic. However, the precise effects of travel restrictions on the outbreak dynamics of COVID-19 remain unknown. Here we combine a global network mobility model with a local epidemiology model to simulate and predict the outbreak dynamics and outbreak control of COVID-19 across Europe. We correlate our mobility model to passenger air travel statistics and calibrate our epidemiology model using the number of reported COVID-19 cases for each country. Our simulations show that mobility networks of air travel can predict the emerging global diffusion pattern of a pandemic at the early stages of the outbreak. Our results suggest that an unconstrained mobility would have significantly accelerated the spreading of COVID-19, especially in Central Europe, Spain, and France. Ultimately, our network epidemiology model can inform political decision making and help identify exit strategies from current travel restrictions and total lockdown.

## 1. Introduction

On March 13, 2020, the World Health Organization declared Europe the epicenter of the coronavirus 2019 pandemic with more reported cases and deaths than the rest of the world combined (World Health Organization 2020). The first official case of COVID-19 in Europe was reported in France on January 24, 2020, followed by Germany and Finland only three and five days later. Within only six weeks, all 27 countries of the European Union were affected, with the last cases reported in Malta, Bulgaria, and Cyprus on March 9, 2020. At this point, there were 13,944 active cases within the European Union and the number of active cases doubled every three to four days (Johns Hopkins University 2020). On March 17, 2020, for the first time in its history, the European Union closed all its external borders to prevent a further spreading of the virus (European Commission 2020). The decision to temporarily restrict all non-essential travel was by no means uncontroversial, although it was very much in line with the mitigation strategies of most of the local governments: Italy had introduced a national lockdown on March 9, Germany had implemented school and border closures starting March 13, Spain followed on March 14, and France on March 16. By 18 March, 2020, more than 250 million people in Europe were in lockdown (Wikipedia 2020). As the economic pressure to identify exit strategies is rising, we ask ourselves, how effective are these measures in mitigating the spreading of the COVID-19 and how risky would it be to lift them?

Obviously, we will never know exactly what would have happened without the drastic political measures of increased border control and massive travel restrictions. One possibility to estimate the effectiveness of travel restrictions is mathematical modeling (Hsu 2020). Mathematical modeling of infectious disease dates back to Daniel Bernoulli in 1760 (Bernoulli 1760) and has been widely used in the epidemiology community since the 1920s (Kermack and McKendrick 1927). The most common approach to model the epidemiology of an infectious disease is to represent the stages of the disease through a number of compartments and introduce constitutive relations that define how individual subpopulations transition between them (Hethcote 2000). A popular compartment model is the SEIR model that represents the timeline of a disease through the interplay of the susceptible, exposed, infectious, and recovered populations (Aron and Schwartz 1984). The transition rates *α* from exposed to infectious and *γ* from infectious to recovered are the inverses of the latent period *A* = 1*/α*, the time during which an individual is exposed but not yet infectious, and the infectious period *C* = 1*/γ*, the time during which an individual can infect others (Li and Muldowney 1994). In theory, both are disease specific parameters, independent of country, region, or city. For COVID-19, depending on the way of reporting, they can range between *A* = 2 to 6 days and *C* = 3 to 18 days (Park et al. 2020; Prem et al. 2020).

The most critical feature of any epidemiology model is the transition from the susceptible to the exposed state. This transition is typically assumed to scale with the size of the susceptible and infectious populations *S* and *I*, and with the contact rate *β*, the inverse of the contact period *B* = 1*/β* between two individuals of these populations (Hethcote 2000). The product of the contact rate and the infectious period, *R*_0_ = *C β* = *C/B*, is called the basic reproduction number (Delamater et al. 2019). It defines how many individuals are infected by a single one individual in an otherwise uninfected, susceptible population (Dietz 1993). Naturally, the basic reproduction number provides valuable insight into the transmissibility of an infectious agent, and its change is a critical measure of behavioral changes and political actions (Fang et al. 2020). Here we leave this parameter free and identify it independently for each country in the European Union (Peirlinck et al. 2020).

Especially during the early stages of an outbreak, mobility can play a critical role in spreading a disease (Balcan et al. 2009). To reduce mobility and mitigate the COVID-19 outbreak, many European countries have followed the guidelines by the European Commission and implemented travel restrictions, closed borders, and prohibited non-citizens from entry (European Commission 2020). This has stimulated an active on-going debate about how strong these restrictions should be and when it would be safe to lift them (Chinazzi et al. 2020). Network modeling of travel-induced disease spreading can play a pivotal role in estimating the global impact of travel restrictions (Colizza et al. 2006). On a global level, a reasonable first estimate for the mobility of a population is passenger air travel (Zlojutro et al. 2019). Air travel statistics have been successfully used to mitigate epidemic outbreaks and prevent the spreading between cities, states, or countries (Pastor et al. 2015). Strikingly, by March 22, 2020, the average passenger air travel in Europe was cut in half, and as of today, April 18, it is reduced by 89% in Germany, 93% in France, 94% in Italy, and 95% in Spain (Eurostat 2020). Here we use a network epidemiology model to explore the effect of travel restrictions within the European Union and simulate the outbreak dynamics and outbreak control of COVID-19 with and without the current travel restrictions in place.

## 2. Materials and Methods

### 2.1 Epidemiology model

We model the epidemiology of the COVID-19 outbreak using an SEIR model with four compartments, the susceptible, exposed, infectious, and recovered populations, governed by a set of ordinary differential equations (Hethcote 2000),

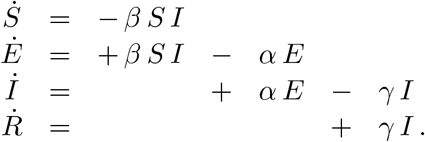

The transition rates between the four compartments, *β, α*, and *γ* are inverses of the contact period *B* = 1*/β*, the latent period *A* = 1*/α*, and the infectious period *C* = 1*/γ*. We interpret the latent and infectious periods *A* and *C* as disease-specific, and the contact period *B* as behavior specific. We discretize the SEIR model in time using an implicit Euler backward scheme and adopt a Newton Raphson method to solve for the daily increments in each compartment.

### 2.2 Mobility model

We model the spreading of COVID-19 through a mobility network of passenger air travel, which we represent as a weighted undirected graph 𝒢 with *N* nodes and *E* edges (Peirlinck et al. 2020). The *N* = 27 nodes represent the countries of the European Union, the *E* = 172 edges the most traveled connections between them. We estimate the mobility within the graph using the annual passenger air travel statistics (Eurostat 2020) from which we create the adjacency matrix, *A*_*IJ*_, that repre-sents the travel frequency between two countries *I* and *J*, and the degree matrix, 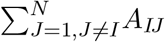, that represents the number of incoming and outgoing passengers for each country *I*. The difference between the degree matrix *D*_*IJ*_ and the adjacency matrix *A*_*IJ*_ defines the weighted graph Laplacian *L*_*IJ*_,

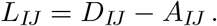

Figure 1, top left, illustrates the discrete graph of the European Union with 27 nodes and 172 edges. The size and color of the nodes represent the degree *D*_*II*_, the thickness of the edges represents the adjacency *A*_*IJ*_. For our passenger travel-weighted graph, the degree ranges from 222 million in Germany, 221 million in Spain, 162 million in France, and 153 million in Italy to just 4 million in Luxembourg, 3 million in Estonia and Slovakia, and 2 million in Slovenia, with a mean degree of 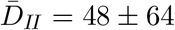 million per node. We assume that the Laplacian *L*_*IJ*_, normalized to one and scaled by the mobility coefficient *ϑ*, characterizes the global spreading of COVID-19. We discretize our SEIR model on our weighted graph 𝒢 and introduce the susceptible, exposed, infectious, and recovered populations *S*_*I*_, *E*_*I*_, *I*_*I*_, and *R*_*I*_ as global unknowns at the *I* = 1, …, *N* nodes of the graph 𝒢. This results in the spatial discretization of the set of equations with 4 *N* unknowns,

**Figure 1.**
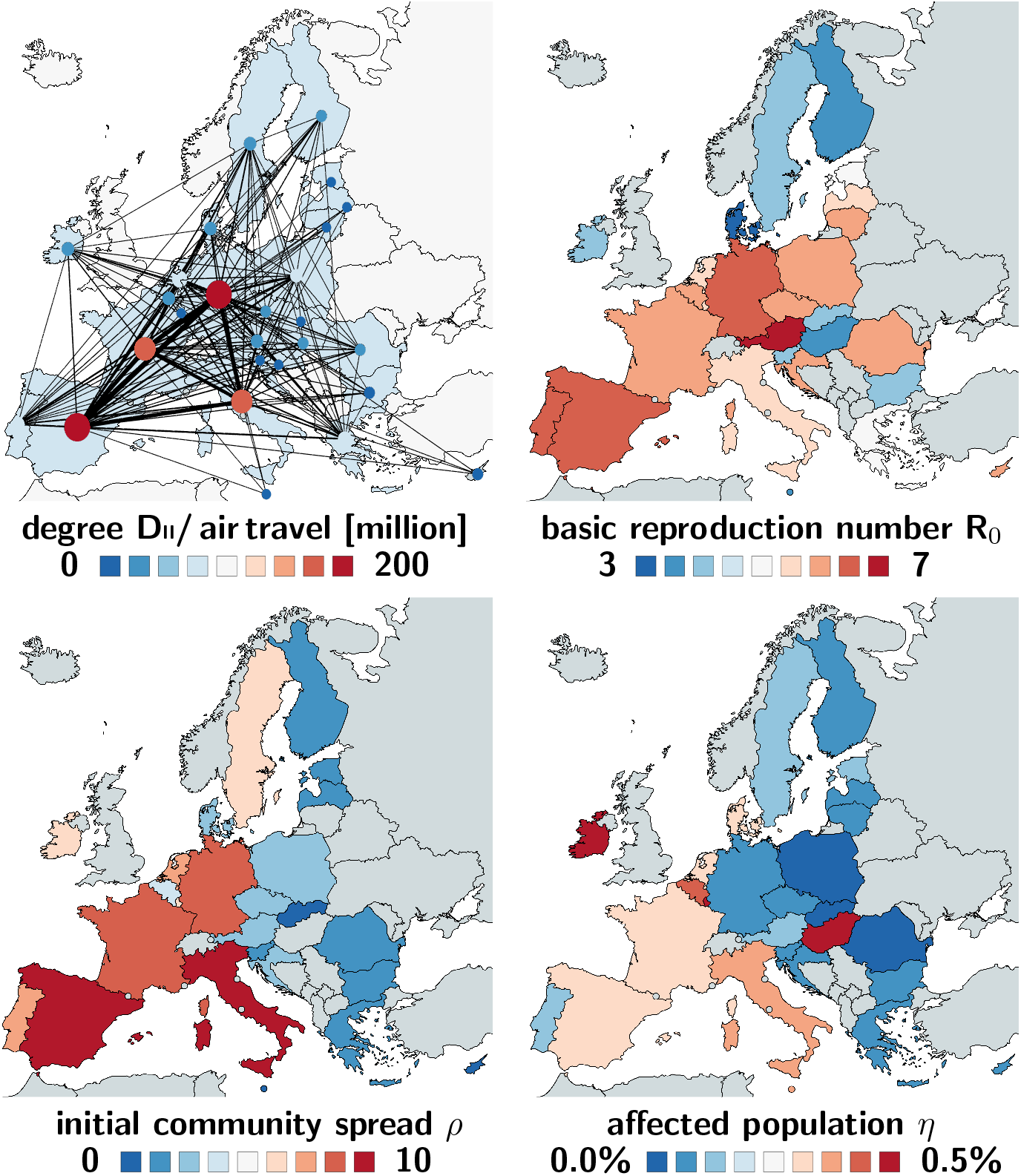
COVID-19 outbreak dynamics across Europe. Mobility network of the European Union with *N* = 27 nodes and the 172 most traveled edges (top left); basic reproduction number *R*_0_ = *C/B* (top right); initial community spread *ρ* = *E*_0_*/I*_0_ (bottom left); and affected population *η* = *N*^*∗*^*/N* (bottom right).

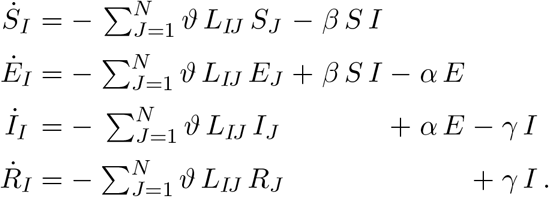

We discretize our SEIR network model in time using an implicit Euler backward scheme and adopt a Newton Raphson method to solve for the daily increments in each compartment in each country (Fornari et al. 2019).

### 2.3 Parameter identification

We draw the COVID-19 outbreak data of all 27 states of the European Union from the temporal evolution of confirmed, recovered, active, and death cases starting January 22, 2020 (Johns Hopkins University 2020). From these data, we map out the temporal evolution of the infectious group *I* as the difference between the confirmed cases minus the recovered cases and deaths. To simulate the country-specific epidemiology of COVID-19 with the SEIR model, we utilize these data to identify the basic reproduction number *R*_0_ = *C/B* using the latent and infectious periods *A* = 1*/α* = 2.56 days and *C* = 1*/γ* = 17.82 days, which we had previously identified for the COVID-19 outbreak in *N* = 30 Chinese provinces (Peirlinck et al. 2020), since the current European data are not yet complete for the identification of all three parameters. In addition to the basic reproduction number *R*_0_ = *C/B*, which is a direct measure of the contact period *B* = 1*/β*, we also identify the initial community spread *ρ* = *E*_0_*/I*_0_ that defines the initial asymptomatic exposed population *E*_0_ (Maier and Brockman 2020), and the affected population *η* = *N*^*∗*^*/N* that defines the fraction of the epidemic subpopulation *N*^*∗*^ relative to the total population *N* (Eurostat 2020). Altogether we identify three parameters for 27 countries, the basic reproduction number *R*_0_ = *C/B*, the initial community spread *ρ* = *E*_0_*/I*_0_, and the affected population *η* = *N*^*∗*^*/N* using the Levenberg-Marquardt method of least squares. For the indentified parameters, we compare two COVID-19 outbreak scenarios: outbreak dynamics with the current travel restrictions, a mobility coefficient of *ϑ* = 0.00, and a country-specific outbreak on day d_0_, the day at which 0.001% of the population is reported as infected; and outbreak dynamics without travel restrictions, with a mobility coefficient of *ϑ* = 0.43 (Peirlinck et al. 2020), and a European outbreak on day d_0_ in Italy, the first country in which 0.001% of the population is reported as infected (Johns Hopkins University 2020).

## 3. Results

Figure 1 shows the basic reproduction number *R*_0_ = *C/B*, the initial community spread *ρ* = *E*_0_*/I*_0_, and the affected population *η* = *N*^*∗*^*/N* across all 27 countries of the European Union. The basic reproduction number is largest in Austria and Germany with *R*_0_ = 8.7 and *R*_0_ = 6.0 and smallest in Malta and Denmark with *R*_0_ = 3.0 and *R*_0_ = 2.7, with a mean of *R*_0_ = 4.62 1.32. The initial community spread is largest in Italy and Spain with *ρ* = 18.4 and *ρ* = 15.2 and smallest in Malta and Cyprus with *ρ* = 0.2 and *ρ* = 0.1 with a mean of *ρ* = 3.53 ± 3.97. The affected population is largest in Ireland and Hungary with *η* = 2.73% and *η* = 1.21% and smallest in Slovakia and Bulgaria with *η* = 0.04% and *η* = 0.02%, with a mean of *η* = 0.08 ± 0.26. Table 1 summarizes the means and standard deviations of our identified parameter values. Figure 2 illustrates the reported infectious and recovered populations and the simulated exposed, infectious, and recovered populations for all 27 countries. The simulations use the basic reproduction number *R*_0_ = *C/B*, the initial community spread *ρ* = *E*_0_*/I*_0_, and the affected population *η* = *N*^*∗*^*/N* identified for each country using disease specific latent and infectious periods of *A* = 2.56 days and *C* = 17.82 days. Day d_0_ indicates the beginning of the outbreak at which 0.001% of the population are infected. The outbreak delay across Europe spans a time window of 24 days, ranging from February 28, 2020 in Italy to March 22, 2020 in Hungary.

**Table 1.** COVID-19 outbreak dynamics across Europe. Latent period *A*, contact period *B*, infectious period *C*, basic reproduction number *R*_0_ = *C/B*, initial community spread *ρ* = *E*_0_*/I*_0_ and affected population *η* = *N*^*∗*^*/N* with means and standard deviations for all 27 countries of the European Union.

| parameter | mean ± std | interpretation |
| --- | --- | --- |
| **A [days]** | $2.56 \pm 0.72$ | **latent period** |
| **B [days]** | $4.07 \pm 1.23$ | **contact period** |
| **C [days]** | $17.82 \pm 2.95$ | **infectious period** |
| **$R_0$ [-]** | $4.62 \pm 1.32$ | **basic reproduction no** |
| **$\rho$ [-]** | $3.53 \pm 3.97$ | **initl latent population** |
| **$\eta$ [-]** | $0.0767 \pm 0.2612$ | **affected population** |

**Figure 2.**
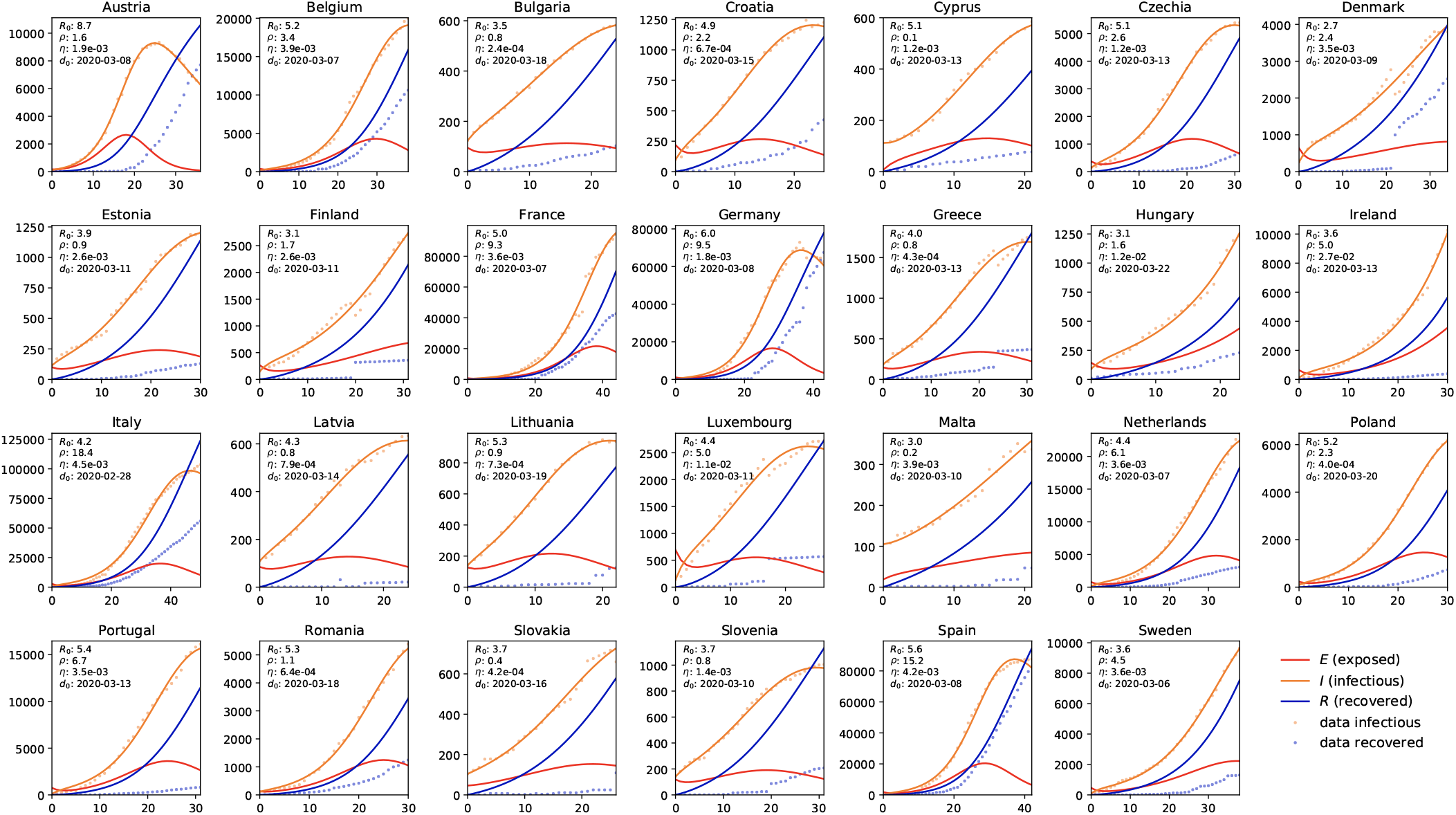
COVID-19 outbreak dynamics across Europe. Reported infectious and recovered populations and simulated exposed, infectious, and recovered populations. Simulations are based on a parameter identification of the basic reproduction number *R*_0_ = *C/B*, the initial community spread *ρ* = *E*_0_*/I*_0_, and the affected population *η* = *N*^*∗*^*/N* for each country for given disease specific latent and infectious periods of *A* = 2.56 days and *C* = 17.82 days. The day d_0_ indicates the beginning of the outbreak at which 0.001% of the population are infected.

Figure 3 highlights the effects of the COVID-19 outbreak control across Europe. The top row shows the simulated outbreak under constrained mobility with the current travel restrictions and border control in place; the bottom row shows the outbreak under unconstrained mobility without travel restrictions. The spreading pattern in the top row follows the number of reported infections; the spreading pattern in the bottom row emerges naturally as a result of the network mobility simulation, spreading rapidly from Italy to Germany, Spain, and France. During the early stages of the pandemic, the predicted outbreak pattern in the bottom row agrees well with the outbreak pattern in the top row. During the later stages, the side-by-side comparison shows a faster spreading of the outbreak under unconstrained mobility with a massive, immediate outbreak in Central Europe and a faster spreading to the eastern and northern countries.

**Figure 3.**
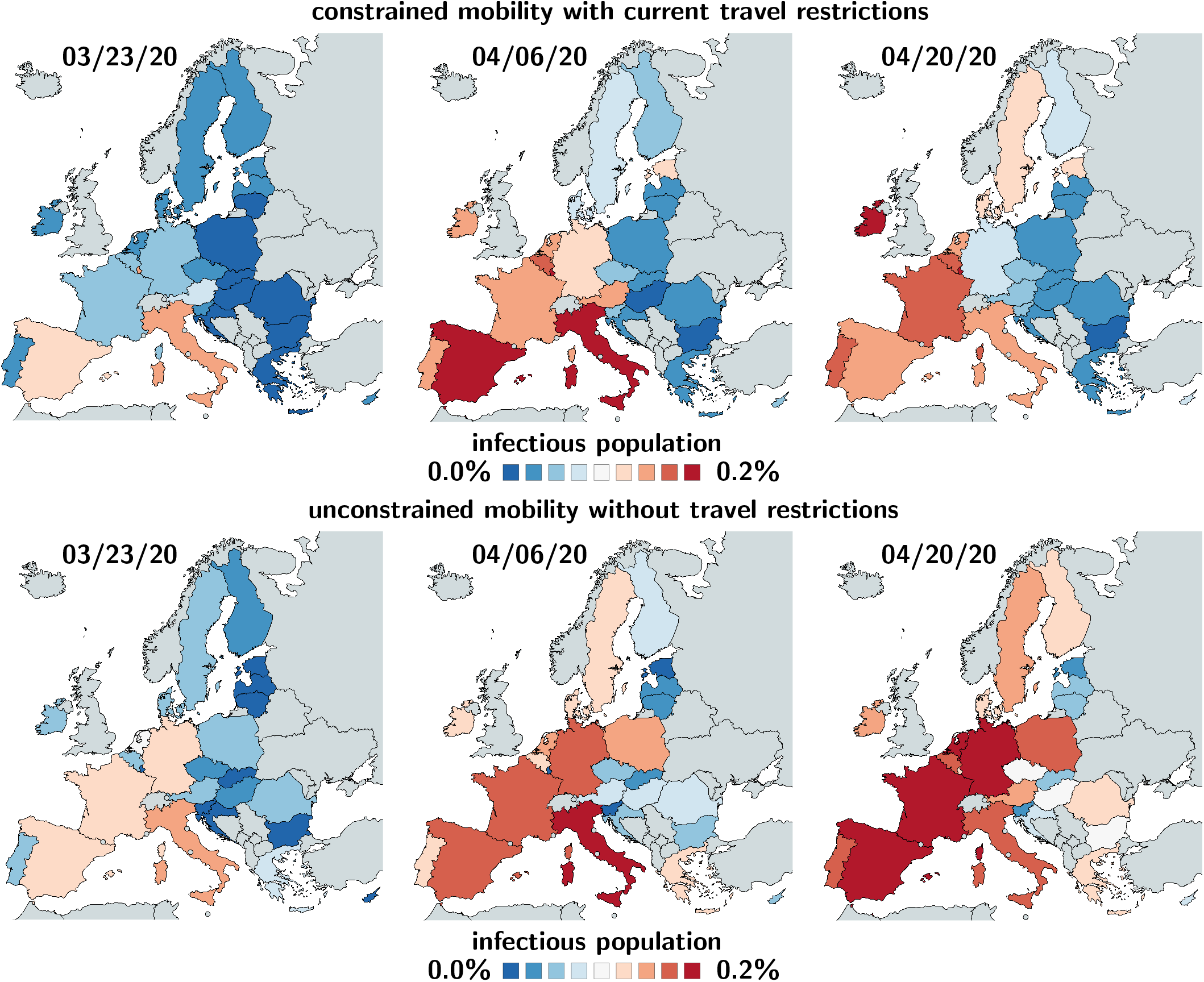
COVID-19 outbreak control across Europe. Effect of travel restrictions. Constrained mobility with travel restrictions (top) vs. unconstrained mobility without travel restrictions (bottom). Simulations are based on latent, contact, and infectious periods of *A* = 2.56 days, *B* = 4.07 days, and *C* = 17.82 days, and mobility coefficients of *ϑ* = 0.00 (top) and *ϑ* = 0.43 (bottom).

## 4. Discussion

Freedom of movement is the cornerstone of the European Union. On March 26, 2020, the 25th anniversary of the Schengen Agreement that guarantees unrestricted movement between its 27 member countries, all external and many internal boarders of the European Union were closed to reduce the spreading of the coronavirus (European Commission 2020). These drastic measures have stimulated a wave of criticism, not only because they violate international law, but also because of a strong belief that they only slow, but do not halter, the spread of the pandemic (Mason Meier et al. 2020). A recent study based on a global metapopulation disease transmission model for the COVID-19 outbreak in China has shown that the Wuhan travel ban essentially came too late, at a point where most Chinese cities had already received many infected travelers (Chinazzi et al. 2020). Our study shows a similar trend for Europe, where travel restrictions were only implemented a week after every country had reported cases of COVID-19 (Johns Hopkins University 2020). Nonetheless, our side-by-side comparison of simulations with and without travel restrictions illustrates the rapid spreading of COVID-19 across Central Europe. Our results suggest that the emerging pattern of the COVID-19 outbreak closely follows global mobility patterns of air passenger travel (Pastor et al. 2015): From its European origin in Italy, the coronavirus spread rapidly via the strongest network connections to Germany, Spain, and France, while slowly reaching the less connected countries, Estonia, Slovakia, and Slovenia.

Figure 3 supports the decision of the European Union and its local governments to implement rigorous travel restrictions in an attempt to delay the outbreak of the pandemic. Austria, for example, rapidly introduced drastic mitigation strategies including strict border control and massive travel bans. Its national air travel was cut in half by March 20 and soon after converged to a reduction of 95% (Eurostat 2020). This has rapidly reduced the number of new cases to a current fraction of 10% of its all time high three weeks ago (Johns Hopkins University 2020), which has motivated a gradual lift of the current constraints. Our prognosis for Austria in Figure 3 suggests that, without travel restrictions, Austria would still see a rise of the infected population. Critics argue that community-based public health measures such as physical distancing, contact tracing, and isolation would be equally successful, but less restrictive, alternatives to restricting the freedom of movement (Mason Meier et al. 2020). Admittedly, this points toward a limitation of our current model in which the basic reproduction number is constant and does not account for local mitigation strategies (Peirlinck et al. 2020). We are currently improving our model to dynamically adapt the reproduction number and correlate its changes to the timing and severity of political actions.

As we are trying to identify exit strategies from local lockdowns and global travel restrictions, political decision makers are turning to mathematical models for quantitative insight and scientific guidance. There is a well-reasoned fear that easing off current measures, even slightly, could trigger a new outbreak and accelerate the spread to an unmanageable degree. Global network mobility models, combined with local epidemiology models, can provide valuable insight into different exit strategies. Our model allows us to virtually lift travel restrictions between individual communities, states, or countries, and explore gradual changes in spreading patterns and outbreak dynamics. Our results demonstrate that mathematical modeling can provide guidelines for political decision making with the ultimate goal to gradually return to normal while keeping the rate of new COVID-19 infections steady and manageable.

## Data Availability

all data are available in public data bases

## Acknowledgments

This work was supported by a Stanford Bio-X IIP seed grant to Mathias Peirlinck and Ellen Kuhl and by a DAAD Fellowship to Kevin Linka.

## References

J. L. Aron, I. B. Schwartz. Seasonality and period-doubling bifurcation in an epidemic model. J. Theor. Bio. 110 (1984) 665–679.

D. Balcan, V. Colizza, B. Goncalves, H. Hu, J. Jamasco, A. Vespignani. Multiscale mobility networks and the spatial spreading of infectious diseases. Proc. Nat. Acad. Sci. 106 (2009) 21484–21489.

D. Bernoulli. Essay d’une nouvelle analyse de la mortalite causee par la petite verole et des avantages de l’inoculation pour la prevenir. Mem. Math. Phys. Acad. Roy. Sci. Paris (1760) 1–45.

M. Chinazzi, J. T Davis, M. Ajelli, C. Gioanni, … A. Vespignani. The effect of travel restrictions on the spread of the 2019 novel coronavirus (COVID-19) outbreak. Science (2020) doi:10.1126/science.aba9757.

V. Colizza, A. Barrat, M. Barthelemy, A. Vespignani. The role of the airline transportation network in the prediction and predictability of global epidemics. Proc. Nat. Acad. Sci. 103 (2006) 2015–2020.

P. L. Delamater, E. J. Street, T. F. Leslie, Y. T. Yang, K. H. Jacobsen. Complexity of the basic reproduction number (R_0_). Emerg. Infect. Disease 25 (2019) 1–4.

K. Dietz. The estimation of the basic reproduction number for infectious diseases. Stat. Meth. Med. Res. 2 (1993) 23–41.

European Commission. COVID-19: Temporary restriction on non-essential travel to the EU. Communication from the Commission to the European Parliament, the European Council and the Council. Brussels, March 16, 2020.

Eurostat. Your key to European statistics. Air transport of passengers. https://ec.europa.eu/eurostat ; assessed: April 12, 2020.

Y. Fang, Y. Nie, M. Penny. Transmission dynamics of the COVID-19 outbreak and effectiveness of government interventions: a data-driven analysis. J. Med. Virol. (2020) 1–15.

S. Fornari, A. Schafer, M. Jucker, A. Goriely, E. Kuhl. Prion-like spreading of Alzheimer’s disease within the brain’s connectome. J. Royal Soc. Interface. 16 (2019) 20190356.

H. W. Hethcote. The mathematics of infectious diseases. SIAM Review 42 (2000) 599–653.

J. Hsu. Here’s how computer models simulate the future spread of new coronavirus. Scientific American (2020) February 23, 2020.

Johns Hopkins University. Coronavirus COVID-19 Global Cases by the Center for Systems Science and Engineering. Baltimore, 2020. https://coronavirus.jhu.edu/map.html, https://github.com/CSSEGISandData/covid-19; assessed: April 4, 2020.

W. O. Kermack, G. McKendrick. Contributions to the mathematical theory of epidemics, Part I. Proc. Roy. Soc. London Ser. A 115 (1927) 700–721.

M. Y. Li, J. S. Muldowney. Global stability for the SEIR model in epidemiology. Math. Biosci. 125 (1984) 155–164.

B. F. Maier, D. Brockmann. Effective containment explains sub-exponential growth in confirmed cases of recent COVID-19 outbreak in mainland China. medRxiv (2020) doi:10.1101/2020.02.18.20024414.

B. Mason Meier, R. Habibi, Y. Tony Yang. Travel restrictions violate international law. Science 367 (2020) 1436.

S. W. Park, B. M. Bolker, D. Champredon, D. J. D. Earn, M. Li, J. S. Weitz, B. T. Grenfell, J. Dushoff. Reconciling early-outbreak estimates of the basic reproductive number and its uncertainty: framework and applications to the novel coronavirus (SARS-CoV-2) outbreak. medRxiv (2020) doi:10.1101/2020.01.30.20019877.

R. Pastor-Satorras, C. Castallano, P. Van Mieghem, A. Vespignani. Epidemic processes in complex networks. Rev. Modern Phys. 87 (2015) 925–979.

M. Peirlinck, K. Linka, F. Sahli Costabal, E. Kuhl. Outbreak dynamics of COVID- 19 in China and the United States. Biomech. Model. Mechanobio. in press (2020) doi:10.1101/2020.04.06.20055863.

K. Prem, Y. Liu, A.J. Kucharski, R.M. Eggo, N. Davies. The effect of control strategies to reduce social mixing on outcomes of the COVID-19 epidemic in Wuhan, China: a modeling study. Lancet Public Health. doi:10.1016/S2468-2667(20)30073-6.

World Health Organization. WHO Director-General’s opening remarks at the media briefing on COVID-19. https://www.who.int/dg/speeches/detail/who-director-general-s-opening-remarks-at-the-mission-briefing-on-covid-19-13-march-2020; recorded: March 13, 2020; accessed: April 18, 2020.

A. Zlojutro, D. Rey, L. Gardner. A decision-support framework to optimize border control for global outbreak mitigation. Sci. Rep. 9 (2019) 2216.

Wikipedia. 2020 Coronavirus pandemic in Europe. https://en.wikipedia.org/wiki/2020 corona-virus pandemic in Europe; accessed: April 18, 2020.

